# Investigating the Role of Vitamin D in the Prevention and Control of Dengue Virus Vectors and Related Diseases: A Systematic Review Study

**DOI:** 10.1101/2025.01.07.25320153

**Authors:** Ebrahim Abbasi

## Abstract

**Introduction:** Dengue fever is one of the most common vector-borne diseases in the world, affecting many people annually and causing many deaths. Besides, treating this disease is difficult, and there is no effective vaccine for it. In recent years, attention has been paid to the role of micronutrients, including vitamin D, in the control and treatment of viral diseases, including dengue fever. Accordingly, this study aimed to investigate the role of vitamin D in the treatment and control of dengue fever worldwide using a systematic review method.

**Methods:** This study was conducted as a systematic review of the role of vitamin D in the prevention and control of dengue fever globally using a systematic review method. Therefore, all relevant articles were extracted and reviewed through a search in the international scientific databases, including PubMed/MEDLINE, WEB OF Science (ISI), and SCOPUS, without a time limit until the end of 2024. The quality of the articles was assessed using the STROB checklist.

**Results:** Six articles published between 2018 and 2023 were included in the systematic review process. According to the findings, vitamin D affects macrophages that are differentiated from monocytes and increases resistance to dengue virus. Vitamin D also reduces pro-inflammatory cytokines, transcription, and reduction of mRNA receptors, increases the production of interleukins, especially IL-10, and plays a role in reducing viral load, severity of clinical symptoms, and infection control.

**Conclusion:** Vitamin D3 can control the disease and decrease viral load and the severity of dengue fever in patients by inhibiting the inflammatory response and enhancing the immune response. However, given the limited number of studies, it is recommended that more studies be conducted in this field so that this can be discussed with more evidence and accuracy.

## INTRODUCTION

Dengue fever is a disease caused by dengue virus (DENV) transmitted by the Aedes aegypti mosquito ^1^. This virus is common in tropical and subtropical regions. Given that more than half of the world’s population lives in these regions, many people are infected with this virus annually. Ninety-six million cases of dengue fever with clinical manifestations and 20,000 deaths occur worldwide annually ^2^. Most cases of dengue fever are asymptomatic and do not require medical care. In acute cases, clinical manifestations include high fever, rash, nausea, vomiting, joint and muscle pain. This disease is divided into three categories based on the severity of symptoms and clinical manifestations: dengue with warning signs (DWWS), severity as dengue without warning symptoms (DNWS), and severe dengue (SD) ^3^. Reinfection with other dengue virus strains leads to exacerbation of clinical manifestations in this disease ^4^.

The Dengvaxia vaccine has been licensed in 20 countries to combat this virus; however, it has not yet been approved as an effective vaccine for widespread immunization ^5, 6^. Given the difficulty of treating this disease and the lack of an effective vaccine, recent attention has been drawn to the use of micronutrients as adjunctive therapy and their role in strengthening and modulating the immune system. These include vitamins A and D, iron, and proteins ^5, 7, 8^.

Micronutrient deficiencies affect host immune system activity; for example, iron deficiency affects T-cell proliferation, phagocyte function, and cytokine activity during pathogenesis ^9, 10^, vitamin A deficiency affects phagocyte numbers and cellular immunity in viral diseases ^11, 12^, and vitamin D deficiency affects phagocytosis, macrophage maturation, cellular immunity, and the synthesis of pro-inflammatory cytokines ^11, 13^. Beyond its role in calcium and phosphorus metabolism, which is essential for bone growth and strength, vitamin D also functions as an immunomodulator, influencing the activity of immune cells such as macrophages, monocytes, and both T and B lymphocytes ^14, 15^. As a result, vitamin D deficiency can impair immune function and be a risk factor for the spread of infection ^16^. Recently, experts have recommended the use of vitamin D to combat viral diseases, including dengue fever. However, the results reported in studies are contradictory. Thus, this study aimed to investigate the effectiveness of vitamin D as an adjunct to the control and treatment of dengue fever worldwide using a systematic review method to achieve a comprehensive result.

## MATERIAL AND METHODS

This study was conducted as a systematic review of the role of vitamin D in the treatment and control of dengue fever worldwide according to the Preferred Reporting Items for Systematic Reviews and Meta-Analysis (PRISMA) guidelines ^17^. This research has been registered in the International Prospective Register of Systematic Review (PROSPERO) with the code CRD42021231605

### 1. Search strategy

Articles were searched in the international databases of PubMed/Medline, Web of Science, and Scopus using the keywords dengue virus, dengue fever, dengue virus infection, DENV-2 infection, dengue hemorrhagic fever, dengue shock syndrome, vitamin D3, vitamin D, and DENV, both individually and in combination, with the use of OR and AND operators. The title, abstract, and full text were searched without a time limit up until the end of 2024, and all relevant articles were retrieved.

### 2. Inclusion and exclusion criteria

Articles with the following criteria were included in the study: 1-A study was conducted on the dengue virus or disease, 2-The use of vitamin D supplementation was investigated, 3-The treatment and clinical outcomes of dengue fever were investigated, 4-The effect of vitamin D supplementation on the disease and virus was investigated, and 6-They were of satisfactory quality. Articles that did not meet the inclusion criteria were excluded.

### 3. Quality assessment

Article quality assessment was performed using the STROBE (Strengthening the Reporting of Observational Studies in Epidemiology) checklist according to the guidelines. The maximum score achievable was 33, and in this study, a score of more than 20 was acceptable ^18^.

### 4. Study Selection

A total of 25,570 articles were extracted in the initial search. Then, the articles were entered into the Endnote software, duplicates were identified, and 1,945 articles were excluded due to duplication. In the next step, by carefully studying the titles and abstracts of the articles, 23,587 articles were excluded due to their irrelevance to the study. Subsequently, the full text of 38 articles was reviewed, of which 32 articles were excluded from the study due to the lack of clear investigation of the type of effect and outcome of vitamin D on dengue fever. Finally, six articles were included in the systematic review process (Figure 1).

**Figure 1.**
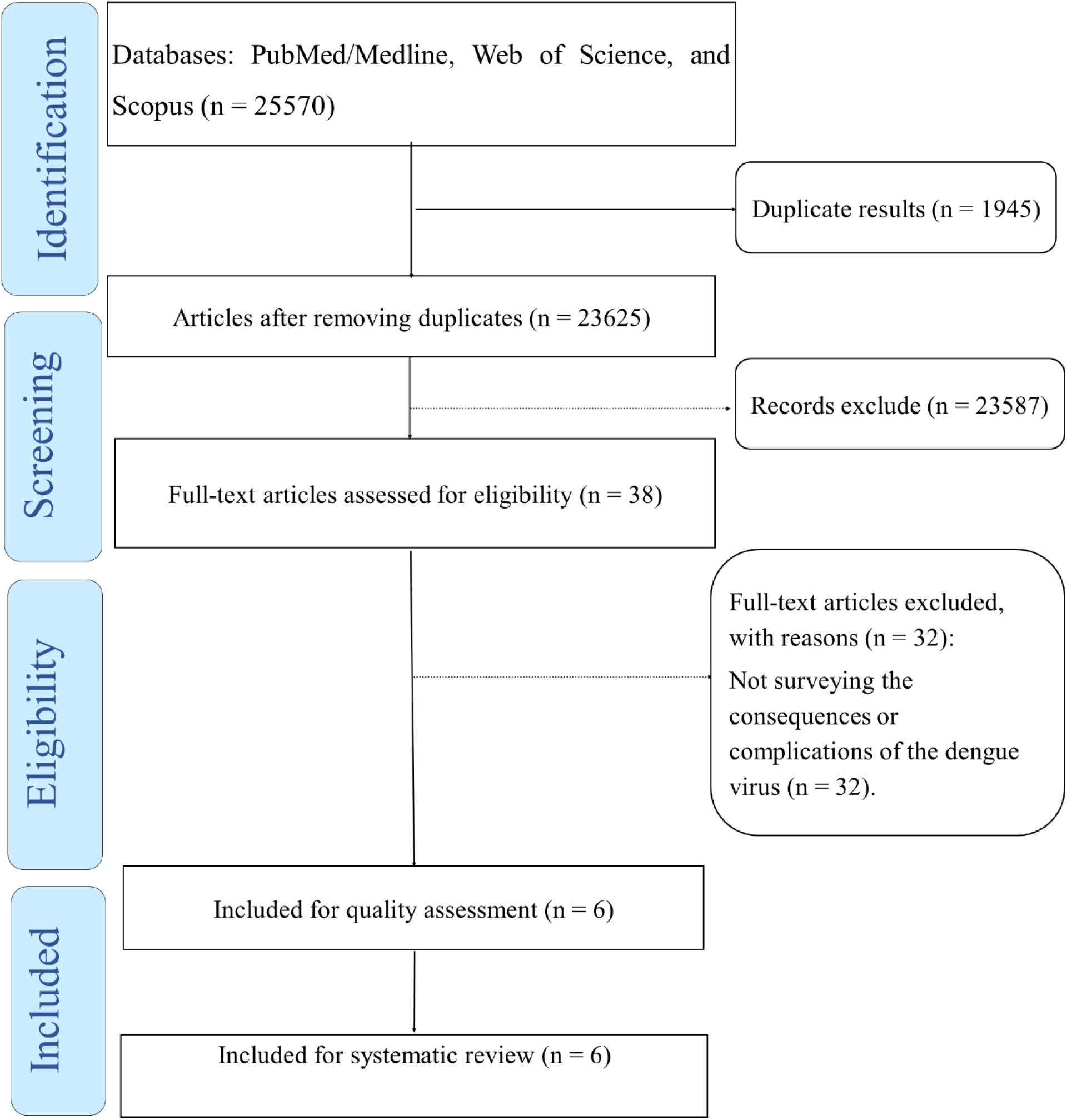
The review process based on the PRISMA flow diagram.

### 5. Data extraction

Data extraction was performed independently by two researchers. Accordingly, the full text of the articles that met the inclusion criteria was first reviewed. If the articles were rejected by the two researchers, the reason was stated, and in case of disagreement between them, the article was reviewed by a third person. Data extraction was performed using a checklist that included the characteristics of the first author, the study author, the publication date of the article, the sample size, the type of intervention, the outcome measured, and the duration of follow-up.

## RESULTS

Six articles that were conducted between 2018 and 2023 were included in the systematic review process. The characteristics of the articles included in the systematic review are presented in Table 1.

**Table 1.** Characteristics of articles included in the systematic review.

| Author | Year of study | Place of study | Exposure | Outcome | Quality assessment |
| --- | --- | --- | --- | --- | --- |
| Castillo JA <sup>21</sup> | 2021 | Colombia | 1α,25 dihydroxy vitamin D3 at a concentration of 0.1 nM | Reducing viral replication and the production of pro-inflammatory cytokines | High |
| **Giraldo DM** <sup>19</sup> | 2018 | Colombia | 1000 or 4000 international units (IU)/day of vitamin D during 10 days | Decreased pro-inflammatory cytokines, intracellular toll-like receptor (TLR), and CAMP mRNA, and increased interleukin 10 (IL-10) production | Mild |
| **Mirza WA** <sup>20</sup> | 2022 | Pakistan | Coinfection H. pylori and dengue fever | Vitamin D deficiency | High |
| **Iqtadar S** <sup>3</sup> | 2023 | Pakistan | Vitamin D deficiency | Severe dengue fever | High |
| **Castillo JA** <sup>23</sup> | 2022 | Colombia | LL-37 | Reduced replication of the virus and production of IL-6 increased the expression of genes involved in virus sensing and antiviral response | High |
| **Castillo JA** <sup>22</sup> | 2023 | Colombia | Vitamin D3 | Expression of inflammatory-liked miR-182-5p, miR-130a-3p, miR125b-5p, miR146a-5p, and miR-155-5p | High |

In a study by Giraldo et al. (2018), monocyte-differentiated macrophages (MDMs) exposed to higher doses of vitamin D (4000 IU/day) showed greater resistance to DENV-2 infection. Also, increasing vitamin D intake in MDMs significantly reduced pro-inflammatory cytokines, intracellular toll-like receptor (TLR), and CAMP mRNA. It increased interleukin 10 (IL-10) production, which may play a role in controlling viral infection. Finally, it was noted that high-dose vitamin D intake could be effective in controlling the progression of dengue fever and DENV replication ^19^. In a study by Mirza et al. (2022) investigating vitamin D deficiency in patients with dengue fever, it was reported that those with co-infection of Helicobacter pylori (which can cause vitamin D deficiency) had a high prevalence of vitamin D deficiency. Also, clinical symptoms of dengue disease, including dizziness, shortness of breath, persistent vomiting, diarrhea, abdominal pain, headache, gingival bleeding, heart rate, and blood pressure fluctuations, were significantly more severe in patients with dengue fever co-infection with H. pylori who were vitamin D deficient ^20^.

In another study, Iqtadar et al. (2023) examined the association between serum vitamin D levels with dengue fever (DF), dengue hemorrhagic fever (DHF), and dengue shock syndrome (DSS). The findings showed that among patients with vitamin D deficiency, 73% had DF, 78.8% had DHF, and 87.5% had DSS, indicating that vitamin D deficiency is associated with dengue fever severity ^3^. Castillo et al. (2021) noted that macrophages, as the main cellular targets for DENV replication, had lower viral replication and produced lower levels of pro-inflammatory cytokines in the presence of vitamin D3. MDMs also expressed lower levels of RIG I, Toll-like receptor (TLR) 3, and TLR7, and higher levels of SOCS-1 against DENV-2 infection in the presence of vitamin D. In general, vitamin D3 modulates the innate immune responses of macrophages by reducing ROS production, downregulating TLRs, and upregulating SOCS 1 and IFN-stimulated genes such as PKR and OAS. Accordingly, they noted that vitamin D3 could have antiviral and anti-inflammatory effects in DENV-2-infected macrophages and could ultimately be a candidate for anti-DENV therapy ^21^. In another study by Castillo et al. (2023), it was shown that inhibition of miR-155-5p, miR-130a-3p, miR-182-5p, and miR-125b-5p resulted in decreased production of TNF-α and TLR9 and increased SOCS-1, IFN-β, and OAS1; however, it did not affect DENV proliferation. Conversely, overexpression of miR-155-5p, miR-130a-3p, miR-182-5p, and miR-125b-5p significantly reduced the infection and proliferation rate of DENV-2 in MDMs. Given that vitamin D3 supplementation differentially regulates the expression of inflammatory microRNAs and can modulate the immune system, vitamin D3 may play a key role in the inflammatory response to DENV infection ^22^. Also, Castillo et al. (2022) showed in another study that simultaneous exposure of LL-37 with DENV-2 during entry into the body leads to a decrease in virus replication in MDMs, but the addition of LL-37 after exposure to DENV-2 has no effect on it. Under conditions of simultaneous exposure, IL-6 production is reduced, and the expression of genes involved in the antiviral response is increased. Considering the low endogenous levels and limited production of LL-37 in MDMs in response to DENV-2 infection, the presence of vitamin D3, which leads to increased differentiation of MDMs, can raise its levels and modulate the strength of the immune system in exposure to DENV ^23^. In general, based on the findings of the present study, vitamin D can play a role in reducing DENV infections, reducing proliferation and burden by affecting immune system mechanisms, and reducing the severity of the disease caused by DENV.

## DISCUSSION

The present study was conducted to investigate the relationship between vitamin D3 and dengue fever virus and disease, and the findings demonstrated that the use of vitamin D3 can be effective in reducing viral replication and in improving and reducing clinical symptoms of patients. Various studies have been performed on the effect of vitamin D3 on viral diseases in the world, indicating that vitamin D3 can be useful in controlling the disease and its clinical symptoms ^24, 25^. Schneider et al. (2014) mentioned that vitamin D in rhinovirus infection increases the secretion of pro-inflammatory chemokines CXCL8 and CXCL10, which play a role in attracting immune cells, including neutrophils, macrophages, and T cells, to the site of infection and induces an antiviral response against HRV infection ^26^. Other studies have shown that vitamin D deficiency is associated with increased susceptibility to respiratory syncytial virus (RSV) in the first year of life of infants ^27^. Treatment with vitamin D, through inhibition or activation of inflammatory markers, increases the level of IκBα, reduces the inflammatory response to RSV infection, increases the antiviral response, and ultimately reduces the severity of complications and mortality from this infection ^28, 29^. Besides, other studies have indicated that vitamin D, as an immunomodulator, plays an important role in inflammatory responses, fibrosis caused by HCV infection, and the development of a persistent viral response ^30^. It also improves the immune response in Peg-α-2b/ribavirin and Peg/RBV treatments ^31, 32^. It indicates that the use of vitamin D supplementation is helpful in the treatment, control, and reduction of the severity and clinical symptoms of viral diseases.

Various mechanisms have been mentioned in the field of the effect of vitamin D on viral diseases. Teymoori Rad et al. (2018) reviewed that vitamin D3 may affect viruses and their associated diseases by inducing an antiviral state, interacting with cellular and viral factors, causing apoptosis and autophagy, genetic and epigenetic changes, functional immunoregulatory properties, and modulating effects on gene transcription ^33^.

Other studies have shown that vitamin D is associated with monocyte function, which is mediated by the CYP27B1 enzyme ^34, 35^. Monocytes use various receptors, including Toll like receptors (TLRs), to recognize foreign bodies and perform phagocytosis. Evidence has shown that CYP27B1 activity is enhanced during this process ^36^. It has also been observed that 1. (OH)2D. 25 is increased during this enhancement and controls gene expression in monocytes ^37^. Following increased gene transcription, it encodes the antibiotic protein LL37 ^38^. Increased LL37 levels lead to improved monocyte function. Finally, it can be noted that vitamin D leads to increased monocyte activity, and vitamin D deficiency can reduce monocyte potency. Other evidence is the role of calcitriol in inhibiting inflammatory T cell cytokines such as IL-2 and IL-17 ^39^. Studies have shown that high doses of calcitriol supplementation in healthy individuals lead to a decrease in the level of the pro-inflammatory cytokine IL-6. The combination of the above-mentioned effects can lead to the induction of regulatory T cells, which are important for regulating immune responses ^40^. So, based on the findings of the present study, it was found that Vitamin D3 can be used as a supplement to treat dengue fever and reduce the severity, clinical symptoms, and viral load.

## CONCLUSION

The present systematic review showed that vitamin D3, through its effect on pro-inflammatory and inflammatory mechanisms, including cytokines, mRNA receptors, and interleukins, can reduce viral load, control disease, and decrease the severity of dengue fever in patients by inhibiting the inflammatory response and enhancing the immune response. However, more studies are required in different regions of the world to discuss this issue with more evidence and accuracy.

## Data Availability

All data produced in the present work are contained in the manuscript

## DECLARATION

### Ethics approval and consent to participate

Not applicable.

### Data Availability Statement

All data generated or analysed during this study are included in this published article.

### Consent for publication

Not applicable

### Authors’ contributions

EA determined the title, wrote and registered the protocol, and submitted the article. EA extracted the files from the databases. EA, screening, and selection of final reports. EA, data extraction. EA wrote the article. All authors read and approved the final manuscript.

## Acknowledgments

The authors thank the Research Vice-chancellor of Shiraz University of Medical Sciences. A preprint has previously been published.

